# Maternal mental health and nutritional status of infants aged under 6 months: a secondary analysis of a cross-sectional survey

**DOI:** 10.1101/2024.04.03.24305269

**Authors:** Mubarek Abera Mengistie, Melkamu Berhane, Carlos S. Grijalva-Eternod, Alemseged Abdissa, Nahom Abate, Endashaw Hailu, Hatty Barthorp, Elizabeth Allen, Marie McGrath, Tsinuel Girma, Jonathan CK Wells, Marko Kerac, Emma Beaumont

## Abstract

Maternal/caregivers’ mental health (MMH) and child nutrition are both poor in low- and middle-income countries. Links between the two are plausible but poorly researched. Our aim was to inform future malnutrition management programmes by better understanding associations between MMH and the nutritional status of infants aged u6m. We conducted a health facility-based cross-sectional survey of 1060 infants in rural Ethiopia, between October 2020 and January 2021. We collected data on: MMH status (exposure) measured using the Patient Health Questionnaire (PHQ-9) and infant anthropometry (outcome); length for age Z-score (LAZ), weight for age Z-score (WAZ), weight for length Z-score (WLZ), mid upper arm circumference (MUAC), head circumference for age Z-score (HCAZ) and lower leg length (LLL). Linear regression analysis was used to determine associations between exposure and outcome variables. Mean (SD) age was 13.4 (6.2) weeks. The median score for MMH problem was 0 (inter quartile range 0 - 2) and 29.5 and 11.2% reported minimal and mild to severe depression score of 1-4 and 5-25, respectively. Mean (SD) LAZ was -0.4 (1.4), WAZ -0.7 (1.3), WLZ -0.5 (1.2), MUAC 12.4 (1.3) centimetre, HCAZ 0.4 (1.3) and LLL 148 (13.9) millimetre. In adjusted analysis, minimal MMH problems was associated with infant LAZ marginally (β=-0.2; 95% CI: -0.4, 0.001) and LLL (β=-2.0; 95% CI: -3.8, -0.1), but not with other anthropometric measurements. Significant associations were not found between mild to severe depressive symptoms and infant anthropometric outcomes. Covariates positively associated with infant anthropometric measurements were higher wealth index with LAZ (β=0.08, 95% CI: 0.03, 0.13), WAZ (β=0.12, 95% CI: 0.08, 0.17), WLZ (β=0.09, 95% CI: 0.05, 0.13), MUAC (β=0.06, 95% CI: 0.02, 0.11), and HCAZ (β=0.07, 95% CI: 0.03, 0.12); higher maternal schooling with LAZ (β=0.24, 95% CI: 0.05, 0.43) and WAZ (β=0.24, 95% CI: 0.07, 0.41); female sex with WAZ (β=0.16, 95% CI: 0.01, 0.31) and HCAZ (β=0.16, 95% CI: 0.001, 0.31); higher maternal age with LLL (β= 0.29, 95% CI: 0.07, 0.52); and improved water, sanitation and hygiene status with MUAC (β=0.07, 95% CI: 0.01, 0.12) and LLL (β=0.64, 95% CI: 0.04, 1.24). Covariates negatively associated with infant anthropometric measurements include female sex with MUAC (β=-0.33, 95% CI: - 0.48, -0.18) and LLL (β=-2.51, 95% CI: -4.15, -0.87); higher household family size with WLZ (β=-0.08, 95% CI: -0.13, -0.02); exclusive breastfeeding with MUAC (β=-0.39, 95% CI: -0.55, - 0.24) and LLL (β=-7.37, 95% CI: -9.01, -5.75); and grandmother family support with WAZ (β=- 0.2, 95% CI: -0.3, -0.0001) and WLZ (β=-0.2, 95% CI: -0.4, 0.1). In conclusion, only minimal, but not mild, moderate or severe, maternal/caregivers’ depressive symptoms are associated with infant anthropometry outcomes. Whilst plausible relationship between maternal mental health problems and offspring nutritional status exist, we are not able to show this because of small number of participants with moderate to severe level of depression in our study population. Thus, further evidence to understand and establish robust relationship between maternal mental health and offspring nutritional status is required.

## INTRODUCTION

Poor level of maternal/caregiver’s mental health (MMH) and poor child growth are major public health problems in low-and middle-income countries (LMICs) (1,2). Poor MMH represents a decline in the status of mental well-being during pregnancy or within a year of childbirth.

Among various MMH problems, depression and anxiety during the pre-and post-natal periods are most common and increasing, particularly in LMICs. Systematic review and meta-analysis evidence showed prevalence of 25% (3) and 33% (4) peri- and post-natal depression, respectively in LMICs. Several factors such as pregnancy and child birth-related changes, role transition during and after pregnancy, intimate partner violence, low maternal social capital and poverty contribute to the high burden of MMH problems in LMICs (1). In most LMICs health care delivery systems, MMH problems are treated within the general mental health care system making the service inaccessible.

The burden of malnutrition/poor growth, manifested as stunting, wasting, and underweight, are highest in children aged under-five years in LMICs (2). Though sometimes treated separately in prevention and treatment programmes, there are common features between these different manifestations of malnutrition, most notably a short-term high mortality risk (5) and long-term risks of poor developmental outcomes and non-communicable diseases (6,7). Stunting for example starts in utero and continues during childhood (8,9). Currently, nearly 150 million children aged under-five years are stunted, mostly in LMICs (10). Stunting prevalence is as high as 50% in some settings (11), 42% in Sub Saharan Africa and 37% in Ethiopia in 2020 (2).

Wasting is another major challenge associated with high case fatality and contributing to some 800,000 child-deaths per year (12). Risk factors for child malnutrition include infections and poor social, economic and environmental factors (13).

Linkages between MMH and child nutritional status are plausible. Some evidence suggest that poor MMH is a risk factor for poor child nutrition and growth (14,15) but such evidence, especially in infants under-6 months (u6m), is scarce and complex (16–19). Some of these studies showed a negative impact of MMH problems on infant nutritional status while other reported a lack of association or presence of complex and indirect relationships between the two. More evidence on this is needed, especially since the new WHO guidelines on malnutrition highlight the need to consider MMH as part of care for infants u6m at risk of poor growth and development (as well as older children(20)). There is increasing interest into how to integrate MMH and malnutrition programming (21) so evidence on this area are essential.

In this study, we hypothesise that poor MMH is associated with poor infant u6m nutritional status. Our aim was to examine associations between MMH problems and infant nutritional status to add new knowledge on existing evidence in a low income setting in order to inform and guide future malnutrition prevention and intervention programmes.

## METHODS AND MATERIALS

### Study design

This is a secondary analysis of a previously reported community-based health facility survey in Ethiopia (22). The survey was done in order to inform planning for a randomised controlled trial (23).

### Setting

The original survey included health facilities in Jimma zone (food secure) and Deder district (food insecure), Ethiopia. The total population of Jimma zone is estimated to be over 3.6 million while that of Deder district is approximately 360,980 people (24). Both sites are predominantly agrarian, and their livelihood is dependent on cash crop production such as coffee and khat followed by cattle rearing. In terms of health care coverage, Jimma zone has about 124 health centres and seven hospitals and Deder district has 8 health centres. The data was collected between 12 October 2020 and 29 January 2021.

### Participants

A consecutive sampling method, over a 2-week period, was employed in each of 18 health centres to recruit a total of 1060 infants u6m. Recruitment was from delivery, immunization, and growth monitoring services and under-five clinics. We included all subjects available in the original dataset. The detailed procedures and main findings of the study on anthropometric outcomes are previously reported (22).

### Measurements

#### Main exposure variable

MMH was assessed using the Patient Health Questionnaire (PHQ-9). Presence of mental health symptoms over the last 14 days on PHQ-9 were scored as symptom presenting 0 = not at all, 1= over several days, 2 = more than half the days, and 3 = nearly every day. The raw sum score of individual PHQ-9 items ranges from 0 to 27. Risk of depression is classified as 0 (none), 1-4 minimal), 5-9 (mild), 10-14 (moderate), 15-19 (moderately severe) and 20-27 (severe) depression (25). These cut offs were used in the statistical analysis.

#### Outcome variables

infant anthropometry/nutritional status (Length for age z-score (LAZ), weight for age z-score (WAZ), weight for length z-score (WLZ), mid-upper arm circumference (MUAC), head circumference for age z-score (HCAZ) and lower leg length (LLL)). Length was measured using the UNICEF length/height board to the nearest completed 0.1 cm. Weight was measured using a digital weight scale (Seca 354) to the nearest 5g if weight < 10kg or to the nearest 10g if weight ≥ 10kg. Measurement procedures followed the WHO Child Growth Standard protocols. All anthropometric indices were measured in duplicate.

#### Household related covariates

level of maternal/caregiver education, household family size, and numbers of dependent children aged <18 years, maternal/caregiver’s age, grandmother family support, household wealth status, and water and sanitation hygiene (WASH) status.

#### Infant related covariates

sex, age and date of birth, birth status (singleton, twin, triplet, etc), birth order, number of siblings and breastfeeding status.

### Data analysis

The data was analysed using Stata (StataCorp. 2021) Statistical Software: Release 17.0 College Station, Texas 77845 USA: StataCorp LLC). Percentage and frequency were reported for categorical data while mean, standard deviation, median and inter quartile range (IQR) are presented for continuous data after checking normality of the distribution. Exclusive breastfeeding was considered if the mother/caregiver reported that the infant was entirely breastfed and was not given anything apart from breastmilk in the last 24 hour.

Anthropometric measurements were calculated as the average of a pair of measurements. Nutritional status for Z-scores were computed using length, weight, age and sex of the infant based on the 2006 WHO Child Growth Standards, using the *zanthro* Stata command. LAZ<-6.0 or >6.0, WAZ <-6.0 or >5.0, WLZ<-5 or >5.0 and length <45cm were considered outlier and excluded from the analysis in the same order from the dataset.

To test for robust relationships between MMH and infant nutritional status, unadjusted and adjusted linear regression analysis were conducted by considering different sets of confounders (infant (age, sex, birth order, and breastfeeding status), maternal (age, educational status and presence of grandmother support) and household characteristics (family size, wealth index, and water, sanitation and hygiene status)).

### Ethics

Ethical approval for the original data was granted by Jimma University Institutional Review Board (ref: IHRPGD/478//2020) and by the Research Ethics Committee at the London School of Hygiene and Tropical Medicine (ref 18022). Written informed consent was obtained from the mother/caregivers (guardians) of every infant for participation. This secondary analysis, looking at mental health in detail, is directly covered by the original protocol and ethics under the secondary objective “to assess the mental health status of mothers and caregivers of surveyed newborns and infants u6m.”

## RESULTS

### Participants’ background

From the total of 1060 participants included in the original dataset, 1036 participants were included in the current analysis. Figure 1 shows study participant flow chart.

**Figure I:**
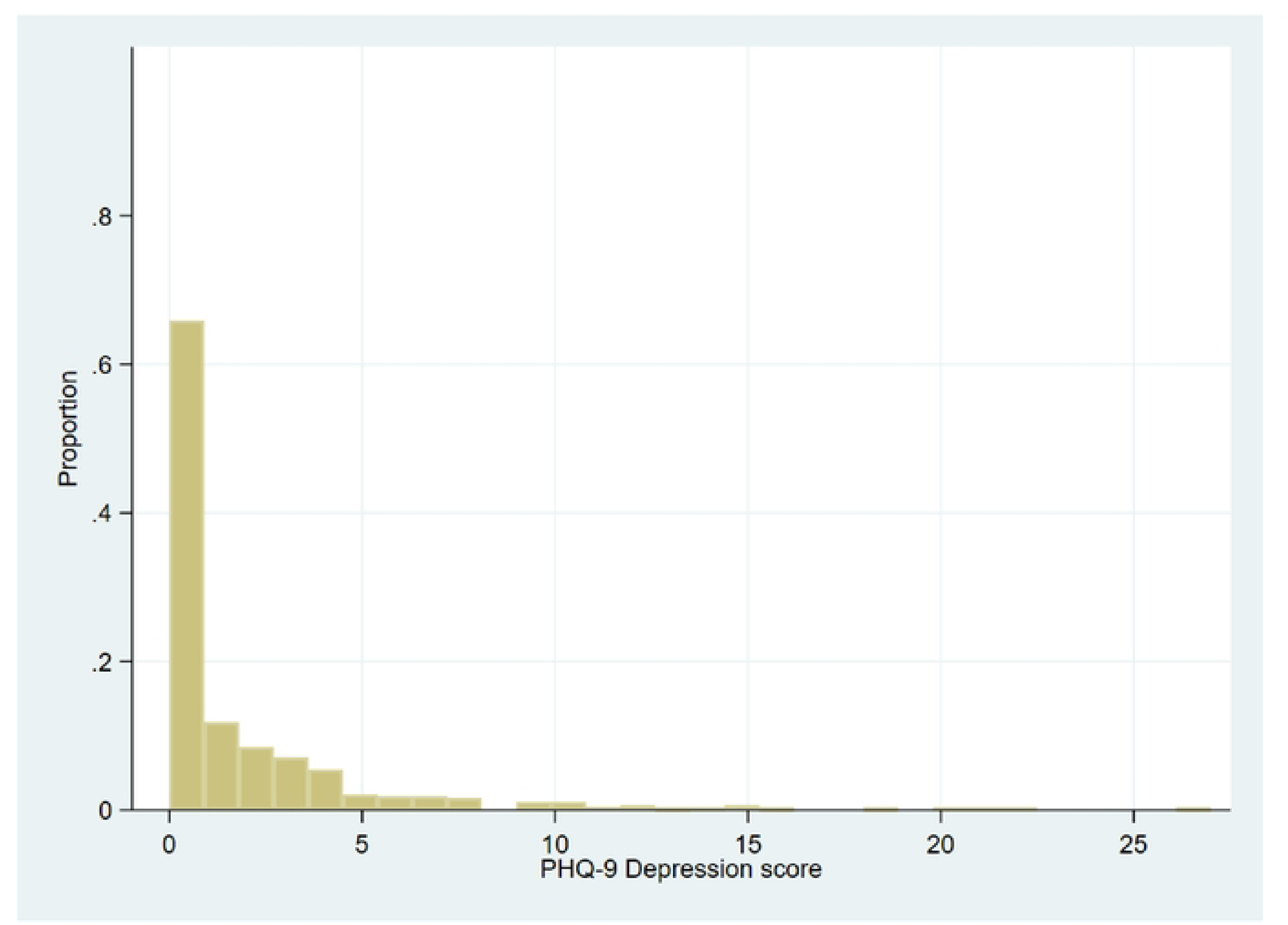
Study participant flow chart

Of those analysed, 577 (55.3%) infants were male and 262 (25.1%) were first borns. Infant’s mean age (SD) was 12.8 (±6.2) weeks. Mean (SD) maternal/caregiver’s age was 25.9 (±5.4) years, and more than half of the mothers/caregivers 619 (59.4%) had attended at least some level of formal education. Nearly 60% of the households had family size of five or less.

### Exposure and outcome characteristics

In terms of MMH, 920 (88.8%) of the mother/caregiver scored none-minimal depression [614 (59.3% no depression and 306 (29.5%) minimal depression], while the rest 116 (11.2%) scored mild to severe depression [78 (7.5%) mild, 23 (2.2%) moderate, 8 (0.8%) moderately severe, and 7 (0.7%) severe depression]. Figure 2 show MMH histogram. The median MMH score was 0 and IQR of 0-2.

**Figure 2:**
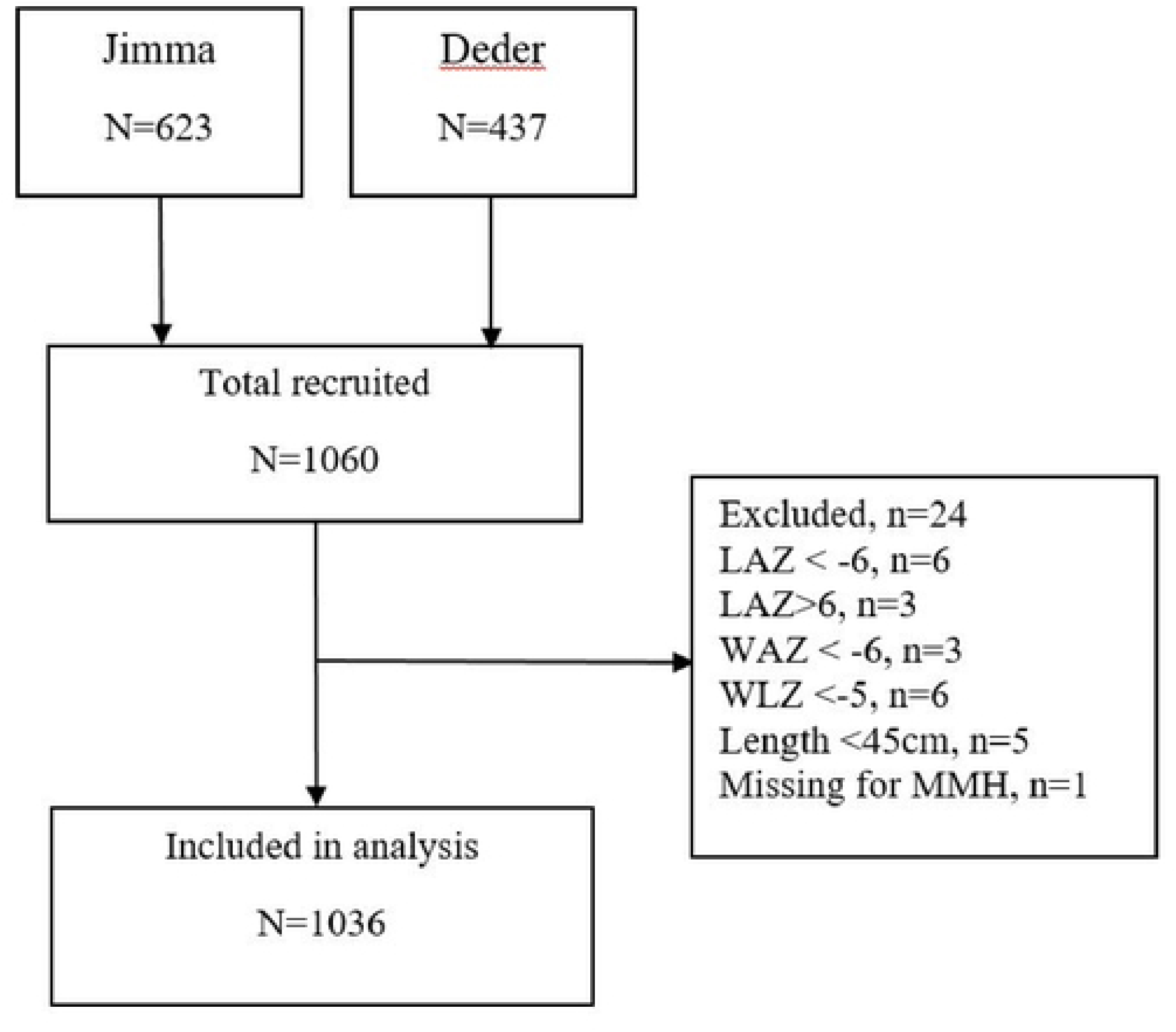
Maternal mental health score histogram

The overall mean and 95% confidence interval (CI) for infant nutritional status was LAZ -0.4 (- 0.4, -0.3), WAZ -0.7 (-0.7, -0.6) and WLZ -0.5 (-0.6, -0.4). All were below the mean value for WHO reference data. The mean (95% CI) value for MUAC was 12.4cm (12.3, 12.5), HCAZ 0.4 (0.3, 0.5) and LLL 148mm (147.1, 148.9). Comparison for infants’ nutritional status by level of maternal mental health is presented in table 1. Figure 3 shows scatter plot between MMH problem score and infant LAZ.

**Figure 3:**
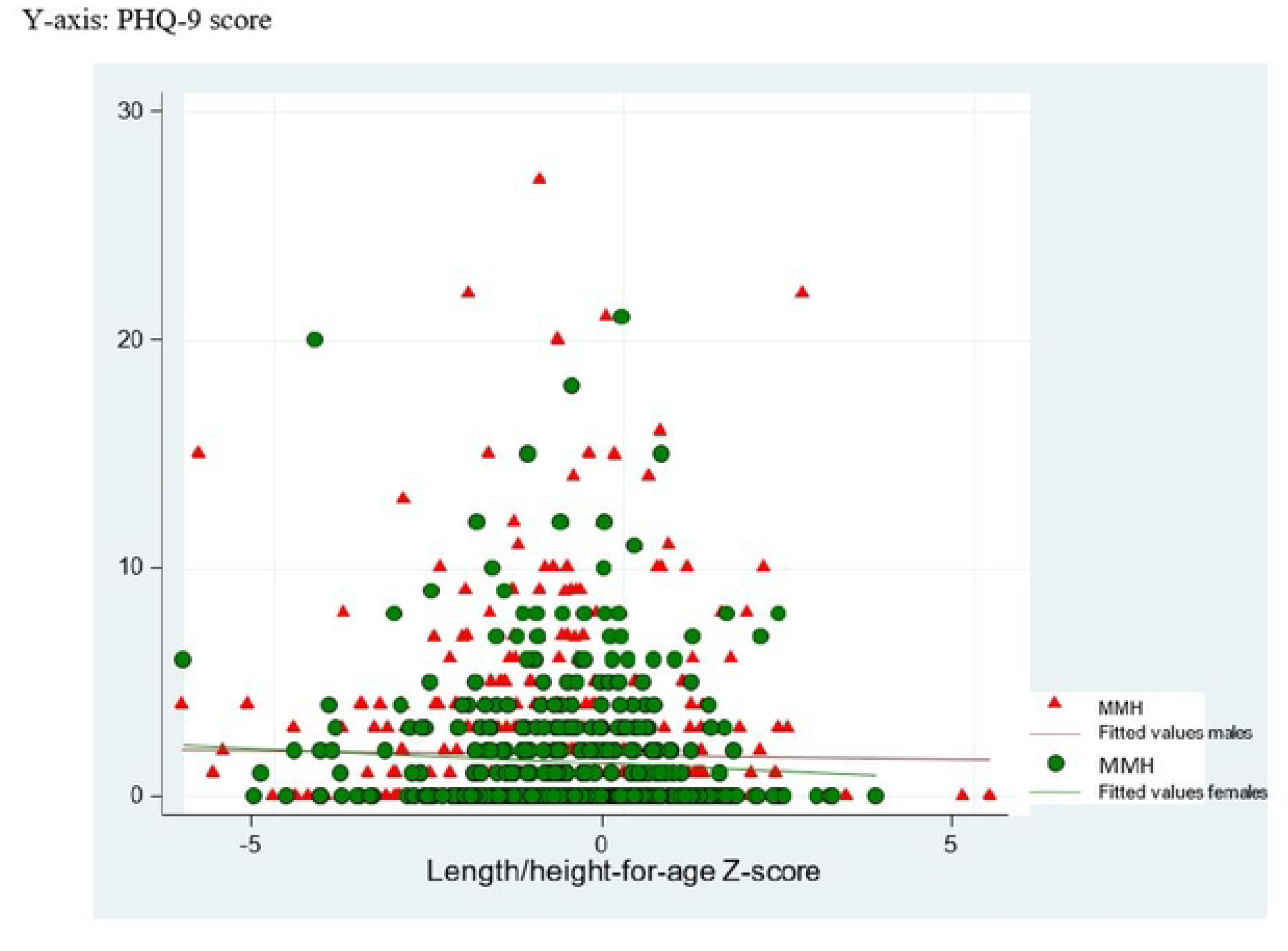
Scatter plot between PHQ-9 score and infant Length for age Z-score

**Table 1:** Background characteristics of study participants, n=1036.

| **Table 1: Background characteristics of study participants, n=1036** |  |  |  |
| --- | --- | --- | --- |
| **Characteristics** | **Category** | **n(%), mean ± SD** |  |
| Infant sex | Male | 574 (55.4) |  |
| Infant age | Weeks | 13.4 ± 6.2 | |
| Infant birth order | 1<sup>st</sup> born | 259 (25.0) |  |
|  | 2<sup>nd</sup> born | 214 (20.7) |  |
|  | 3<sup>rd</sup> born | 156 (15.1) |  |
|  | 4<sup>th</sup> born | 127 (12.3) |  |
|  | 5<sup>th</sup> born | 116 (11.2) |  |
|  | >5<sup>th</sup> born | 164 (15.8) |  |
| Primary caregiver's age, years | | 25.9 ± 5.4 | |
| Primary caregiver schooling | Ever attended school | 612 (59.1) |  |
| Household size | 1-4 | 441 (42.6) |  |
|  | 5-6 | 306 (29.5) |  |
|  | >6 | 289 (27.9) |  |
| Maternal/caregivers mental health | PHQ score 0 | 614 (59.3) |  |
|  | PHQ score 1-4 | 306 (29.5) |  |
|  | PHQ score 5-9 | 78 (7.5) |  |
|  | PHQ score 10-14 | 23 (2.2) |  |
|  | PHQ score 15-19 | 8 (0.8) |  |
|  | PHQ score 20-27 | 7 (0.7) |  |
| Infant nutritional status | All participants (N=1036) | Poor maternal mental health (PHQ 5-27) (n=116) | Maternal mental health (PHQ 0-5) (n=920) |
| Length for age Z-score | -0.35 ± 1.4 | -.25 ± 1.25 | -.35 ± 1.37 |
| Weight for age Z-score | -0.65 ± 1.3 | -.65 ± 1.27 | -.65 ± 1.27 |
| Weight for length for age Z-score | -0.48 ± 1.2 | -.59 ± 1.22 | -.47 ± 1.24 |
| MUAC, centimetre | 12.4 ± 1.3 | 12.31 ± 1.34 | 12.45 ± 1.25 |
| Head circumference Z-score | 0.41 ± 1.3 | .52 ± 1.42 | .40 ± 1.25 |
| Lower leg length, millimetre | 148.07 ± 13.92 | 149.07 ± 16.54 | 147.06 ± 13.6 |

### Association between MMH and infant nutritional status

In Table 2 we reported associations of MMH and infant anthropometric measurements. In unadjusted statistical analysis, only minimal depression (PHQ score 1-4) compared to no depression (PHQ score 0) was negatively associated with LAZ (β=-0.2; 95% CI: -0.4, -0.01) and LLL (β=-2.0, 95% CI: -3.9, -0.1), but not with other anthropometric measurements: WAZ (β=- 0.1; 95% CI: -0.3, 0.04), WLZ (β=0.02; 95% CI: -0.2, 0.2), MUAC (β=-0.1, 95% CI: -0.3, 0.1) and head circumference (β=-0.1, 95% CI: -0.3, 0.1). In the final adjusted statistical analysis, minimal maternal depression was associated negatively with LAZ (β=-0.2; 95% CI: -0.4, 0.0) marginally and LLL (β=-2.0; 95% CI: -3.9, -0.1) and PHQ score 1-27 was associated with LLL ((β=-1.8; 95% CI: -3.4, -0.1). In both unadjusted and adjusted statistical analysis, significant associations were not found between mild, moderate, moderately severe, and severe level of depression with any of infant anthropometric measurements (LAZ, WAZ, WLZ, MUAC, HCAZ and LLL).

**Table 2:**
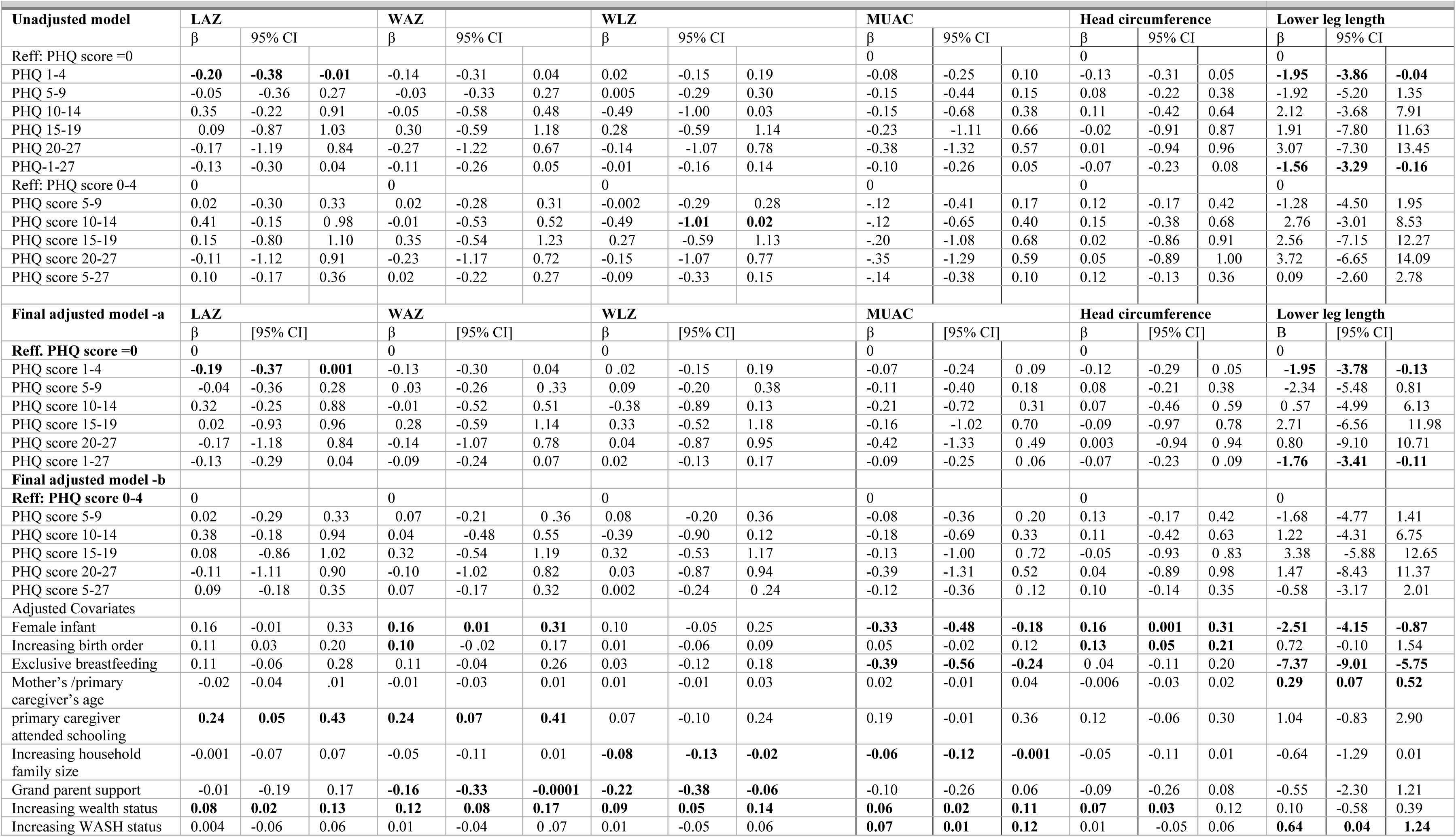
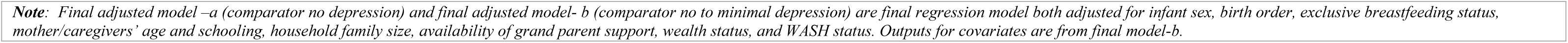
Associations between MMH and infant nutritional status in unadjusted and adjusted statistical models, n=1036.

Covariates positively associated with infant anthropometric measurements were higher wealth index with LAZ (β=0.08, 95% CI: 0.03, 0.13), WAZ (β=0.12, 95% CI: 0.08, 0.17), WLZ (β=0.09, 95% CI: 0.05, 0.13), MUAC (β=0.06, 95% CI: 0.02, 0.11), and HCAZ (β=0.07, 95% CI: 0.03, 0.12); higher maternal schooling with LAZ (β=0.24, 95% CI: 0.05, 0.43) and WAZ (β=0.24, 95% CI: 0.07, 0.41); female sex with WAZ (β=0.16, 95% CI: 0.01, 0.31) and HCAZ (β=0.16, 95% CI: 0.001, 0.31); higher maternal age with LLL (β= 0.29, 95% CI: 0.07, 0.52); and improved water, sanitation and hygiene status with MUAC (β=0.07, 95% CI: 0.01, 0.12) and LLL (β=0.64, 95% CI: 0.04, 1.24).

Covariates negatively associated with infant anthropometric measurements include female sex with MUAC (β=-0.33, 95% CI: -0.48, -0.18) and LLL (β=-2.51, 95% CI: -4.15, -0.87); higher household family size with WLZ (β=-0.08, 95% CI: -0.13, -0.02); exclusive breastfeeding with MUAC (β=-0.39, 95% CI: -0.55, -0.24) and LLL (β=-7.37, 95% CI: -9.01, -5.75); and grandmother family support with WAZ (β=-0.2, 95% CI: -0.3, -0.0001) and WLZ (β=-0.2, 95% CI: -0.4, 0.1).

### Discussion

In this study we examined associations between maternal/caregiver’s mental health with infant nutritional status as indicated by anthropometry. The prevalence of maternal depressive symptoms (PHQ score 5-27) is only 11.2% and the mean (SD) nutritional status of infants u6m was LAZ -0.4 (1.4), WAZ -0.7 (1.3), WLZ -0.5 (1.2), MUAC 12.4 cm (1.3), HCAZ 0.4 (1.3) and LLL is 148 mm (14.0). The prevalence of maternal depressive symptom is lower than previous reports including from other studies in Ethiopia (26,27). Mild, moderate, moderately severe and severe depressive symptoms showed no association with any of the nutritional status indicators. Only minimal depressive symptom associated with low LAZ marginally and low LLL but not with other anthropometric measurements.

The lack of associations between higher maternal depressive symptoms with infant nutritional status is consistent with previous studies where maternal common mental problems were not associated with nutritional status of under five years children in Ethiopia (14,28,29), but contrasts with several other studies from LMICs (16–19,30–35) including findings from Ethiopia (36,37). Our finding is consistent with findings of high income settings (European cohort study (38) and the generation R study (39)) and upper middle income countries (e.g. Brazil) (40) where higher maternal depressive symptoms showed no association with children nutritional status (38–40). In general, associations between postnatal MMH problems and poor child nutritional status have been mainly found in LMICs while lack of associations are mainly from high income setting (15). Unlike our study, previous studies did not included tests of associations between minimal depressive symptoms and childhood nutritional status. Furthermore, the intensity of relationship and direction of associations between these two major public health problems are not well established.

Previously reported associations between maternal mental health problems and poor childhood nutritional status (16–19,30–35) can be explained by several causal mechanisms involving poor maternal child care practice and episodes of childhood illness (4,15,41). Maternal depressive symptoms associated with poor infant feeding practices (42), poor infant breastfeeding outcomes including shorter duration of exclusive breastfeeding (15), poor mother-to-infant bonding (15), low oxytocin total T4 concentration (43) and low breastmilk secretory immunoglobulin A concentration (44) in the breastmilk. Children especially in their early life are entirely dependent on their mother/caregiver’s competencies and resources for nutrition, survival, growth and development. Maternal postnatal depression affects maternal ability and competences to provide responsive and supportive care to her baby (15). This could result in poor maternal-child attachment, bonding, breastfeeding and stimulation; all of which could affect offspring nutrition and growth. Moreover, depressed mothers have poor emotional sensitivity and responsiveness to their environment including their babies (15) which may put at risk infant without support.

Nurturing care as proposed by UNICEF and WHO for child growth and development requires 5 domains of care (good health, adequate nutrition, safety and security, responsive care and early opportunity for learning) (45). All of these domains require good maternal mental health and competencies to manage holistic aspects of her child’s condition.

However these causal pathways through which MMH problems impact offspring nutritional status and our proposed hypothesis are not supported in the current findings. Moreover the lack of association between MMH problems and poor child nutritional status in the previous three studies from Ethiopia (14,28,29) and from upper middle income and high income countries were (38–40) not adequately discussed. Several factors such as the setting and context how the data was collected, types of screening tool used, and the skills, competencies and confidence of data collectors in asking sensitive questions in mental health could have all played a role for the lack of association between MMH problems and poor child nutritional status.

### Implication of the finings

Despite plausible relation between MMH problems and poor infant/child nutritional status exist, our data do not support this. The lack of association in the current study by itself is not evidence of an absent link between the two problems. Moreover, the lack of association between severe MMH problems and infants nutritional status in the current study may be explained by the low prevalence of moderate to severe depression; though previous studies from Ethiopia had failed to establish association despite high prevalence of maternal depressive symptoms were reported.

Depressive disorder by its nature is unique in its clinical presentation compared to many other psychiatric illness in that most patient with depression have good level of insight and understanding about their ill mental health status. This may lead to under or over report based on the context and circumstances how and where the screening process takes place. The high prevalence of minimal depressive symptoms over the low prevalence of moderate to severe depression may indicate that majority of severely affected participants could minimize symptoms due to lack of confidence to report symptoms in a non-conducive screening environment.

Screening for depression requires an established rapport and therapeutic relationship which could take longer time or repeated visits to encourage clients to report their problem by resisting stigma and discrimination. These could have implication to future integration of MMH care in to the maternal and child health service which are too crowded to ensure adequate assessment, privacy and confidentiality. While there is increasing interest into how to integrate MMH and malnutrition programming (21), screening strategy for MMH in the management of small and nutritionally at-risk infants aged under 6 months and their mothers (MAMI) context may require a unique rather than the routine mental health screening approach to be able to effectively elicit symptoms. In general additional qualitative, epidemiological and experimental studies are essential to understand various circumstances that could explain the current finings to reach on robust conclusion.

### Strengths and limitations

We have focused on young infants (u6m) where there is a lack of evidence on this area to support clear decision making process. We covered two different geographic settings representing food secure and insecure areas. Moreover we have included large number of study participants. This study however is not without limitations. The study is a facility based survey and therefore may not be representative of the general population. Moreover the setting where MMH data were collected within the maternal and child health unit may not be conducive enough to establish trusted relationship between participants and study nurses. Finally, though PQH-9 has been widely used in LMIC settings, including in Ethiopia, it may not be optimal in identifying MMH symptoms in the MAMI context and other screening tools/strategies may be more valid and may have shown different association with anthropometric deficit.

## CONCLUSIONS

Mild to moderate and severe levels of maternal/caregiver’s mental health problems are not associated with poor infant nutritional status. Whilst there is plausible relationship between MMH problems and offspring nutritional status, this hypothesis is not supported in the current study. Thus, further evidence to understand and establish the relationship between postnatal maternal mental health problems and infant u6m nutritional status is required both in Ethiopia and other LMIC settings.

## Data Availability

All data are available with the manuscript

